# Comparative Analysis of the Carbon Footprint of Biologics for Severe Asthma

**DOI:** 10.1101/2025.08.08.25333119

**Authors:** Haroon Taylor, Nazneen Rahman

## Abstract

**Objective:** To quantify, compare, and analyse the cradle-to-gate carbon emissions of biologic treatments for severe asthma.

**Design:** Cradle-to-gate carbon emissions for six monoclonal antibody therapies were calculated using MCF Classifier. A representative patient, eligible for all therapies, was defined to enable comparisons. Sensitivity, scenario, and pairwise analyses were conducted to explore variation in emissions and opportunities for reduction.

**Outcome Measures:** The primary outcome was first-year carbon emissions of biologic treatments for a representative patient with severe asthma, expressed in kg CO_2_e. Additional outcome measures were the effect of varying electricity source on treatment emissions, and the emissions associated with alternative biologic choices.

**Results:** First-year treatment emissions for the representative patient ranged from 1.1 kg CO_2_e with benralizumab to 188.9 kg CO_2_e with dupilumab, a 172-fold difference. Variation was driven by the active pharmaceutical ingredient per preparation (30-300 mg). The number of preparations required for first-year treatment (8-52) and the manufacturer’s proportion of non-fossil fuel electricity (NFFE) (22-91%). Sensitivity analysis showed that increasing NFFE to 100% would reduce emissions by 29-90% and the difference between the highest and lowest emission treatments by 91%. Pairwise comparison showed that selecting any biologic instead of dupilumab would reduce emissions by 134-188 kg CO_2_e per patient-year, equivalent to 340-478 car miles. The emission differences between treatment with benralizumab, mepolizumab, and tezepelumab were minimal.

**Conclusions:** The carbon footprints of biologic treatments for severe asthma vary widely, driven primarily by differences in dose and manufacturer electricity sources. MCF Classifier enabled standardised comparisons between therapies and highlighted opportunities for near-term reduction, including increased use of non-fossil fuel electricity and optimisation of dosing practice. The approach can be applied across other therapeutic areas to support carbon-informed prescribing and healthcare decarbonisation.

**Strengths and Limitations of this study:**

- We developed and applied a standardised methodology to estimate cradle-to-gate carbon emissions of biologic treatments for severe asthma, enabling direct comparison across therapies.
- A representative patient was used to ensure consistent application of treatment regimens for each therapy.
- The methodology relies on secondary data and manufacturer-level disclosures, as product-specific primary data is not available.
- The scope was limited to cradle-to-gate emissions, downstream emissions and other components of the clinical pathway were not included.

## Introduction

Asthma affects over 300 million people worldwide and is a leading cause of morbidity.^1^ Although most patients achieve control with inhaled corticosteroids and bronchodilators, a subset experience severe asthma that remains uncontrolled despite high-dose treatment.^2^ These individuals face increased risks of exacerbations, hospitalisation, and long-term lung function decline, and they account for a disproportionate share of healthcare use and cost.^3,4^

Biologic therapies have transformed the management of severe asthma. Six monoclonal antibody treatments are currently licensed for use: benralizumab (Fasenra), dupilumab (Dupixent), mepolizumab (Nucala), omalizumab (Xolair), reslizumab (Cinqair), and tezepelumab (Tezspire). These agents target distinct inflammatory pathways, including interleukin-5 (IL-5) and its receptor, IL-4/IL-13 signalling, Immunoglobulin E, and thymic stromal lymphopoietin (TSLP). Treatment is guided by clinical and biomarker-based phenotyping and has been associated with improved symptom control, reduced exacerbation rates, and lower oral corticosteroid use.^5^

Climate change is recognised as a major determinant of respiratory health.^6^ Rising temperatures, increased air pollution, and extreme weather events contribute to the incidence and severity of asthma, particularly in vulnerable populations.^6^ Healthcare itself is a notable contributor to climate change, responsible for an estimated 4-5% of global greenhouse gas emissions.^7^

The environmental impact of asthma treatment is under increasing scrutiny. Pressurised metered-dose inhalers, which contain hydrofluorocarbon propellants, have high global warming potential, which has led to policy and manufacturing shifts towards lower-emission alternatives. In contrast, the carbon emissions of biologic therapies have received little attention, despite their manufacturing complexity and growing use. A streamlined life cycle analysis estimated emissions exceeding 20,000 kg CO_2_e per kg of product for monoclonal antibodies produced using the US energy grid.^8^ No published data are available on the carbon footprint of individual biologics used in asthma care.

We previously developed and published MCF Classifier, a mass-based, data-driven methodology that integrates modelled data, empirical values, and product-specific attributes to estimate medicine carbon footprints.^9^ The MCF Classifier framework for small molecule medicines is described in Taylor et al.^9^

Here, we extend MCF Classifier to monoclonal antibody biologics and apply it to a comparative analysis of the six biologic therapies currently available for severe asthma.

## Methods

### Biologic selection

We evaluated six monoclonal antibody therapies licensed for the treatment of severe asthma: benralizumab, dupilumab, mepolizumab, omalizumab, reslizumab, and tezepelumab. All were approved in the US and EU at the time of analysis (December 2024). These products were selected to encompass the full range of available biologic treatment options for severe asthma. Where ordering was not determined by analysis, products are listed alphabetically.

### Carbon footprint estimations

We estimated cradle-to-gate carbon emissions for each product preparation using MCF Classifier, a standardised framework for calculating medicine carbon footprints that uses a consistent mass-based approach to enable comparability across products.^9^ We previously published the small-molecule MCF Classifier methodology.^9^ For this study, we applied the biologics module, developed to reflect the specific processes and emission drivers associated with monoclonal antibody production.

We attributed emissions to three components: active pharmaceutical ingredient (API) manufacture and formulation, excipient manufacture, and primary packaging. We excluded downstream activities, including distribution, storage, administration, and disposal.

We followed the GHG Protocol Product Life Cycle Accounting and Reporting Standard and ensured compatibility with ISO 14067 and PAS 2050.^10–12^ A template product carbon footprint report is provided in the Supplementary material.

#### Active Pharmaceutical Ingredient (API)

We estimated the carbon footprint of API manufacturing using a mass-based model that incorporates variation in energy source. We used the global warming potential values over 100 years (GWP) from Budzinski et al. (2022), who estimated emissions for monoclonal antibody production under different electricity grid mixes.^8^ Their values are consistent with those reported in other studies.^13,14^ We modelled the relationship between GWP and non-fossil fuel electricity (NFFE) proportion; with NFFE defined as the proportion (0-1) of electricity from renewable and nuclear sources.

To construct the regression, we took GWP values from Budzinski et al. together with the NFFE proportions from the IRENA energy profiles for the US, China, France, and a 100% wind-powered scenario.^8,15^ The data implied a logarithmic relationship, with greater reductions in GWP at lower NFFE values. We excluded the France data point, as it produced implausible estimates at high NFFE values. Using the remaining values, we derived the following equation:

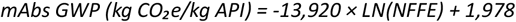

This model was applied for NFFE values between 0.12 and 1.0; estimates below this range were not used.

We obtained manufacturer-level NFFE values from published sustainability disclosures available at the time of the analysis (Table 1).^16–21^ For products with two manufacturers, we used the NFFE value for the API manufacturer. For Genentech, we used the reported NFFE for its parent company, Roche. Company-wide NFFE values were used because neither site-specific nor product-specific data were available. The NFFE values were entered into the regression to calculate a product-specific GWP per kg API, which was then scaled to the API content per preparation.

**Table 1.** Manufacturer non-fossil fuel electricity (NFFE) proportions used in API carbon footprint estimates

| Biologic | Brand name | API Manufacturer | NFFE (%) | Reference |
| --- | --- | --- | --- | --- |
| Benralizumab | Fasenra | AstraZeneca | 91 | [16] |
| Dupilumab | Dupixent | Regeneron | 22 | [17] |
| Mepolizumab | Nucala | GlaxoSmithKline | 83 | [18] |
| Omalizumab | Xolair | Genentech | 81 | [19] |
| Reslizumab | Cinqair | Teva | 43 | [20] |
| Tezepelumab | Tezspire | Amgen | 90 | [21] |

#### Excipients

We estimated excipient emissions using mass-based calculations informed by literature-derived emission factors. Water, typically the largest excipient by volume, was considered to have negligible emissions.^22^ For stabilisers, buffers, and surfactants, we used an estimated emission factor of 5 kg CO_2_e/kg product for microbial fermentation-derived excipients. These excipients accounted for <1% of the total preparation footprint and were treated as negligible.

#### Primary packaging

We estimated packaging emissions using data from Eckelman and Litan.^22^ For pre-filled syringes, we applied cradle-to-gate emission factors based on manufacturing and fill-finish stages of the relevant components (barrel, plunger stopper, staked needle, rigid cover), giving a footprint of 39.8 gCO_2_e per 1 ml syringe. We assumed cradle-to-gate emissions scales linearly with syringe volume for small-volume pre-filled syringes. For vials, we used emissions data for the manufacturing and fill-finish stages of the vial body, crimp, stopper, and sterilization processes, giving 4.7 gCO_2_e per ml. Syringes used for administration of vial-based products were excluded, as they fall outside the cradle-to-gate boundary.

### Representative patient

To enable consistent comparison across biologics with differing eligibility criteria and dosing regimens, we developed a representative patient, referred to as R.P. This allowed standardised application of licensed dosing recommendations for each product to a fixed clinical scenario.

We identified eligibility criteria for each of the six licensed biologics from the FDA and EMA approvals labels.^23–34^ To construct the representative patient, we selected overlapping features that would confer eligibility for each product. These included a diagnosis of severe asthma uncontrolled on high-dose inhaled corticosteroids and additional controller therapies, a recent history of exacerbations, elevated blood eosinophils, elevated IgE, and evidence of perennial allergen sensitivity.

### First-year treatment emissions

We calculated the number of product preparations required to treat R.P. over the first year, based on recommended dosing schedules, including loading doses where applicable, and the presentation format of each product. For products requiring weight-based dosing, we applied a body weight of 70 kg. Preparations were defined as individual units of the relevant format (e.g. a single pre-filled syringe or vial). We multiplied the number of preparations by the carbon footprint per preparation to calculate the total first-year emissions for treating R.P. with each biologic.

### NFFE sensitivity analysis

We conducted a one-way sensitivity analysis to assess how variation in electricity source affects the carbon emissions of R.P.’s first year of treatment with each biologic. Using the regression model described, we varied the NFFE proportion from 0.20 to 1.00 in increments of 0.05, recalculating the GWP of API manufacturing at each point while holding all other model inputs constant. This analysis was performed for each product individually and visualised as a sensitivity curve of first-year emissions versus NFFE proportion.

We then compared baseline first-year emissions with a scenario in which all manufacturers achieved 100% NFFE by recalculating API manufacturing emissions assuming NFFE = 1.00 in the regression model, with all other inputs unchanged. The resulting changes in treatment-level emissions were visualised using a tornado plot.

### Pairwise emissions analysis

We compared first-year emissions between therapies by subtracting the total emissions of a comparator from those of a reference treatment:

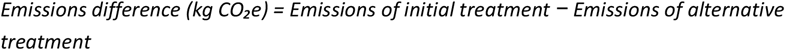

We applied this calculation across all six biologics, to generate a pairwise matrix of first-year emission differences. Negative values indicated higher emissions for the comparator treatment. We presented the results as a heat map. Selected values were also expressed as equivalent car miles using a conversion factor of 0.393 kg CO_2_e per mile.^35^

## Results

### Representative Patient

#### Patient Profile

We defined a standardised representative patient (R.P.) to enable comparison of treatment requirements and associated carbon emissions across biologics with different eligibility criteria and dosing regimens. R.P. was defined based on overlapping eligibility criteria as described in the Methods. The full clinical history is presented in Box 1. All products evaluated were indicated for use in this clinical scenario.

##### BOX 1. Representative Patient (R.P.) Clinical History

R.P. is a 45-year-old female school teacher, of height 165 cm and weight 70 kg. She has severe, persistent asthma with type 2 inflammation, characterised by elevated eosinophil levels and IgE-mediated allergies. Despite adherence to high-dose inhaled corticosteroids and long-acting beta-agonists, she experiences daily asthma symptoms with frequent reliance on a rescue inhaler.

Her asthma is poorly controlled, with five severe exacerbations in the past year requiring oral corticosteroid bursts. She is maintained on oral prednisone 5 mg to help manage exacerbations.

Clinical evaluations revealed an eosinophil count of 450 cells/µL, total serum IgE levels of 400 IU/mL, and confirmed sensitivities to house dust mites and pet dander through allergy testing. Spirometry showed an FEV1 at 60% of predicted, with reversibility exceeding 12% following bronchodilator administration.

**Summary**: R.P. has severe, persistent, uncontrolled asthma despite maximal standard therapy, with type 2 inflammation and allergic sensitivities. She is eligible for respiratory biologic therapies.

#### Treatment recommendations

We determined the recommended treatment schedule for R.P. for each biologic based on the FDA- and EMA-approved labels.^23–34^ For each medicine, we identified the dose, frequency, and form of administration required for the first year of therapy. Loading doses and weight-based adjustments were included, where applicable. The recommendations are summarised in Table 2.

**Table 2.** Biologic treatment regimens and preparations needed for the first year

| Biologic | Treatment regimen for R.P. | Preparations needed for first year |
| --- | --- | --- |
| Benralizumab | 30 mg every 4 weeks for 3 doses, then 30 mg every 8 weeks | 8 x 30 mg |
| Dupilumab | 600 mg loading dose, then 300 mg every 2 weeks | 27 x 300 mg |
| Mepolizumab | 100 mg every 4 weeks | 13 x 100 mg |
| Omalizumab | 225 mg every 2 weeks | 26 x 75 mg + 26 x 150 mg |
| Reslizumab | 210 mg every 4 weeks | 39 x 100 mg |
| Tezepelumab | 210 mg every 4 weeks | 13 x 210 mg |

#### First-year dosing requirements

From the treatment recommendations, we calculated the number of product preparations required for the first year of treatment for each biologic. A preparation was defined as a single unit of the product in its administered form (e.g. one pre-filled syringe or vial). The total number of preparations per year varied from 8 to 52, reflecting differences in dosing frequency, preparation formats, and loading regimens (Table 2).

### Carbon Emissions

#### Product carbon footprints

We estimated the cradle-to-gate carbon footprint for each required preparation. Five biologics required a single preparation per dose. For omalizumab, R.P.’s recommended dose of 225 mg required one 75 mg vial and one 150 mg vial, per dose.

The per-preparation carbon footprints ranged from 139 g CO_2_e to 6,996 g CO_2_e, a 50-fold difference (Table 3). The principal drivers of this variation were the amount of API per dose (30-300 mg) and the manufacturer’s non-fossil fuel electricity proportion (22%-91%; Table 1). Across all products, the API accounted for the majority of emissions, contributing between 71%-99%. Packaging contributed the remainder; excipients were negligible (Table 3).

**Table 3.** Product carbon footprints and the percentage contribution of API and packaging

| Biologic | Product | Carbon footprint |  |  |
| --- | --- | --- | --- | --- |
|  |  | Total (g CO<sub>2</sub>e) | API (%) | Packaging (%) |
| Benralizumab | Fasenra 30 mg, 1 ml pfs* | 138.5 | 71.3 | 28.7 |
| Dupilumab | Dupixent 300 mg, 2 ml pfs | 6996.0 | 98.9 | 1.1 |
| Mepolizumab | Nucala 100 mg, 1 ml pfs | 497.0 | 92.0 | 8.0 |
| Omalizumab | Xolair 75 mg, 0.5 ml pfs | 388.2 | 94.9 | 5.1 |
|  | Xolair 150 mg, 1 ml pfs | 776.5 | 94.9 | 5.1 |
| Reslizumab | Cinqair 100 mg, 10 ml vial | 1419.6 | 96.7 | 3.3 |
| Tezepelumab | Tezspire 210 mg, 1.91 ml pfs | 799.4 | 90.5 | 9.5 |

\*pfs = pre-filled syringe

#### Carbon emissions for first year of treatment

We calculated the total cradle-to-gate carbon emissions for R.P.’s first year of treatment with each biologic by multiplying the number of preparations required by the carbon footprint of each individual preparation. Total emissions ranged from 1.1 kg CO_2_e for benralizumab to 188.9 kg CO_2_e for dupilumab; a 172-fold difference (Figure 1).

**Figure 1.**
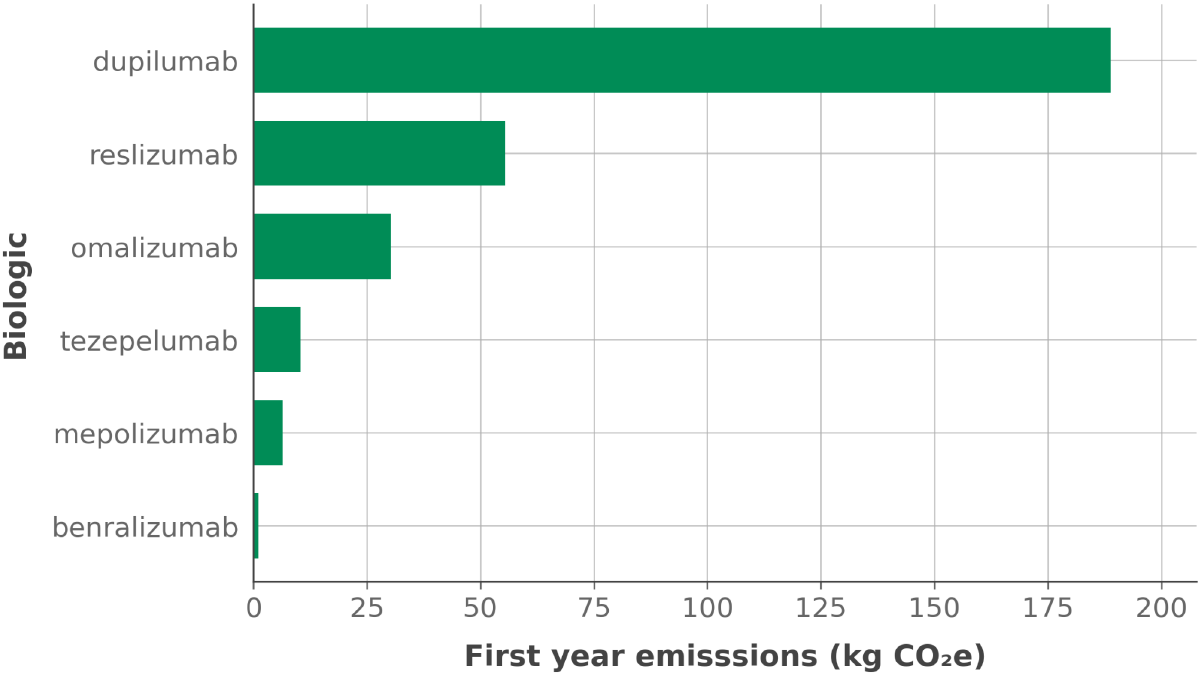
Carbon emissions for R.P.’s first year of treatment with each biologic. Figure legend: Bar chart showing cradle-to-gate emissions (kg CO_2_e) for the representative patient’s first year of treatment with six biologics.

### Sensitivity analysis

We assessed the effect of electricity sourcing on treatment emissions by varying the non-fossil fuel electricity (NFFE) proportion from 0.20 to 1.00. For all products, higher NFFE values reduced the carbon footprint of API manufacture, with the steepest reductions occurring at lower baseline levels of NFFE (Figure 2).

**Figure 2.**
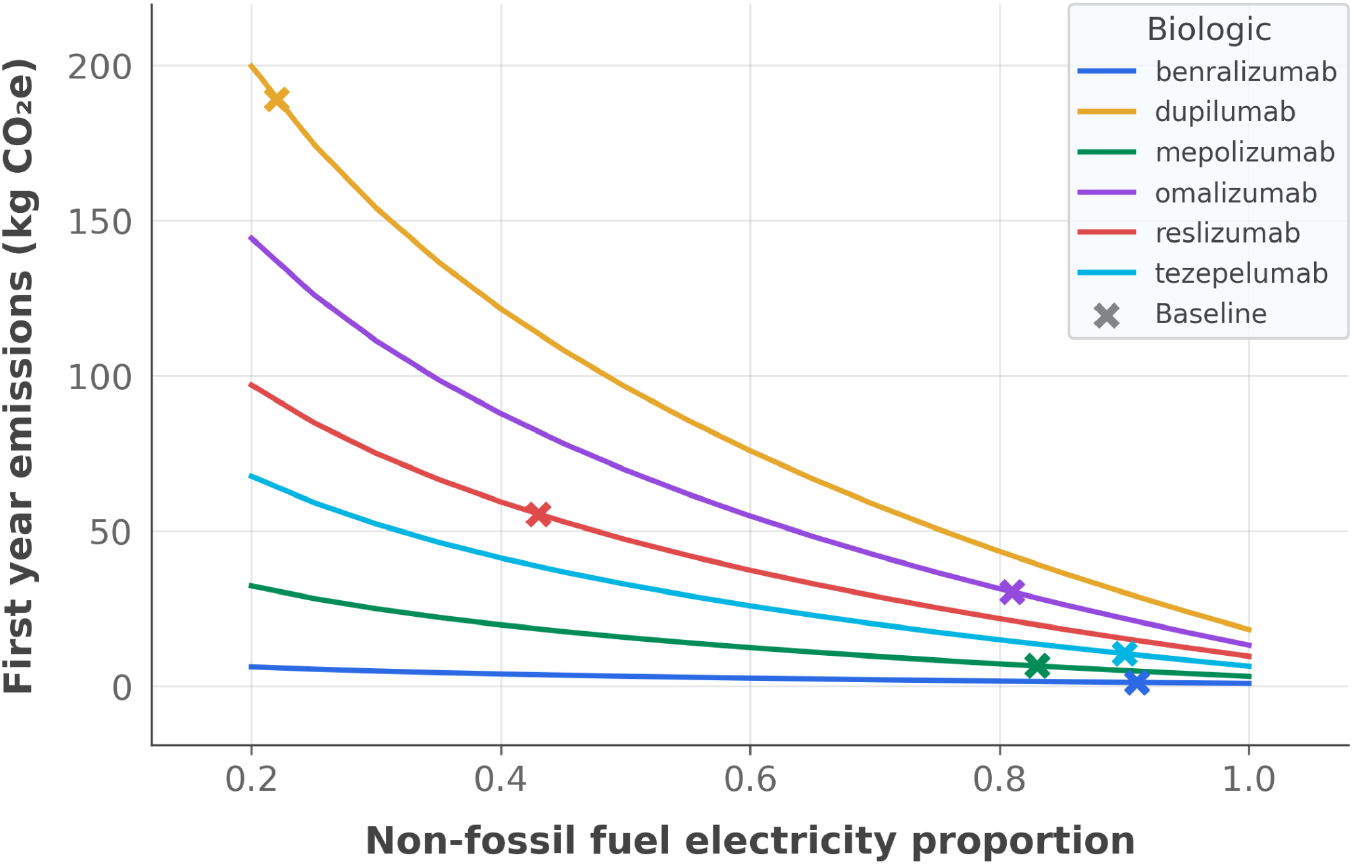
Sensitivity analysis of first-year treatment emissions by NFFE proportion. Figure legend: Sensitivity analysis of first-year treatment emissions by NFFE proportion. Baseline NFFE for each biologic is shown by a cross. Note: “Baseline” refers to the NFFE proportions in Table 1.

We next compared baseline emissions with a scenario in which all manufacturers achieved 100% NFFE. Under this scenario, first-year treatment emissions decreased for all products ranging from a 29% reduction for benralizumab (baseline NFFE 91%) to a 90% reduction for dupilumab (baseline NFFE 22%) (Figure 3). Absolute reductions ranged from 0.3 kg CO_2_e for benralizumab to 170.7 kg CO_2_e for dupilumab, and the fold-difference between the highest and lowest emission treatments fell from 172-fold to 23-fold at 100% NFFE. While most treatments retained their relative ranking, omalizumab replaced reslizumab as the first-year treatment regimen with the second-highest emissions (Figure 3).

**Figure 3.**
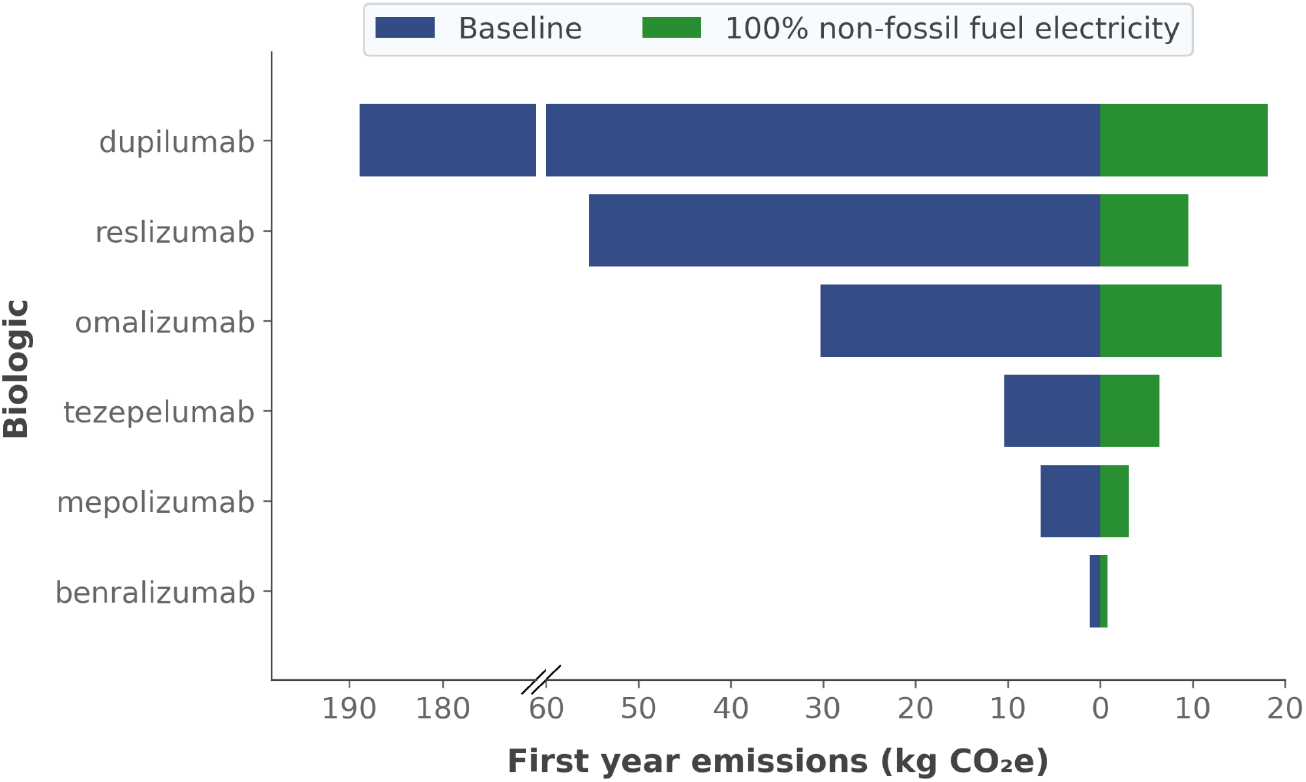
Scenario analysis of first-year treatment emissions at 100% non-fossil fuel electricity. Figure legend: Tornado plot comparing first-year treatment emissions for R.P. under baseline manufacturer-reported non-fossil fuel electricity (NFFE) proportions (blue bars, left) and under a 100% NFFE scenario (green bars, right). Baseline NFFE values are shown in Table 1. A break in the X-axis has been inserted to accommodate the higher emissions associated with dupilumab without compressing the scale for other products.

### Pairwise treatment comparisons

We performed a pairwise analysis to explore the emissions impact of changing treatments. Switching from dupilumab to any other biologic produced the largest reductions in first-year treatment emissions, ranging from 134 kg CO_2_e to 188 kg CO_2_e (equivalent to 340-478 car miles) (Figure 4). In contrast, benralizumab, mepolizumab, and tezepelumab have broadly similar first-year footprints and switches between these three result in only single-digit changes in emissions. Reslizumab and omalizumab occupy an intermediate position; switching either to benralizumab, mepolizumab, or tezepelumab is associated with double-digit reductions in per patient annual emissions.

**Figure 4.**
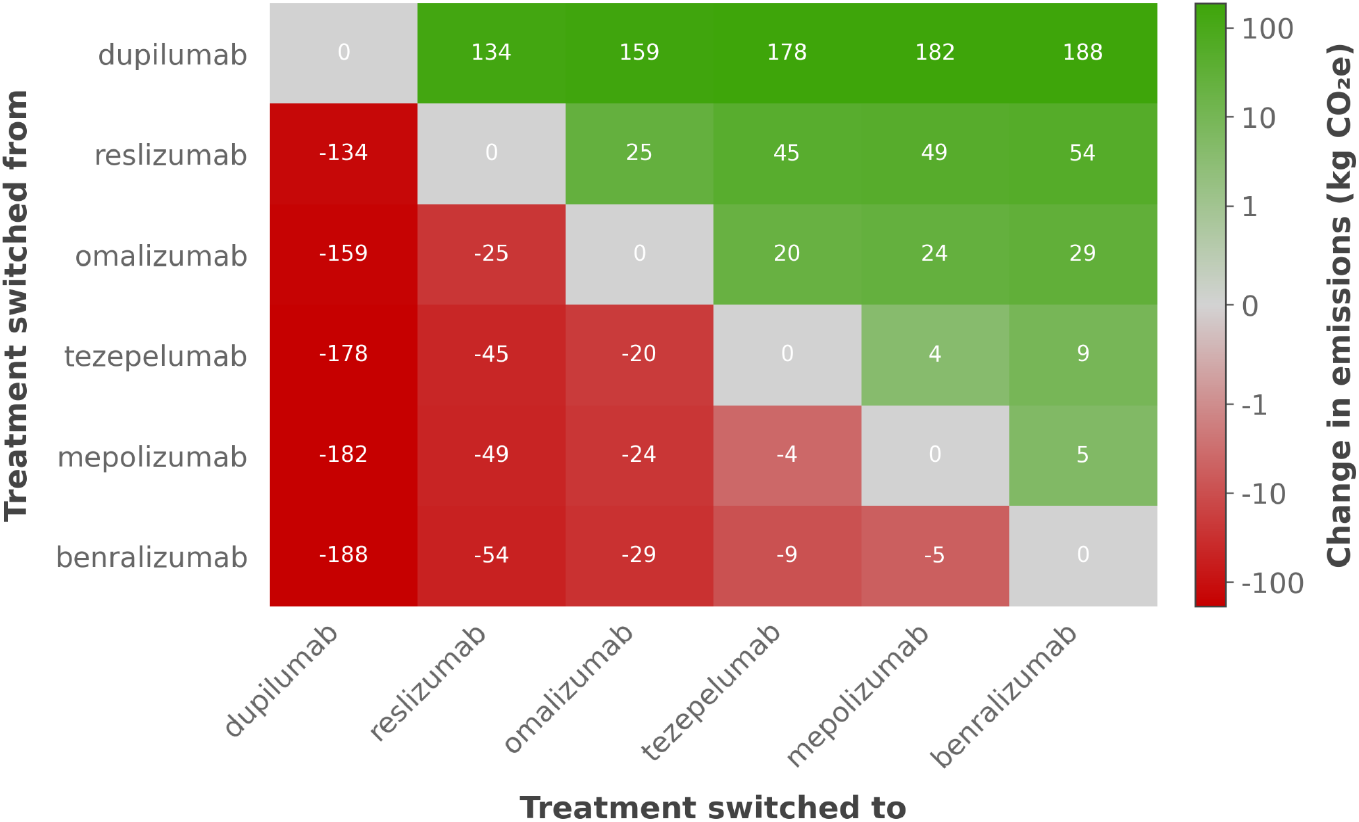
Pairwise comparison of first-year emissions between biologics. Figure legend: Heat map showing the difference in first-year emissions between different biologics. Values represent the change in emissions (kg CO_2_e) relative to the reference treatment in each pairwise comparison. Positive values (green) indicate a reduction in emissions with the comparator treatment; negative values (red) indicate an increase.

## Discussion

Biologic therapies are increasingly used in severe asthma, and our analyses show wide variation in treatment regimens, dosing requirements, and associated carbon emissions. Annual preparation counts ranged from 8 to 52, while the amount of active pharmaceutical ingredient per dose varied from 30 mg to 300 mg. These factors, together with differences in manufacturer electricity profiles, resulted in first-year treatment emissions ranging from 1.1 kg CO_2_e for benralizumab to 188.9 kg CO_2_e for dupilumab. This variation highlights both the scale of emissions associated with biologic treatment and the opportunities to reduce them.

Preparation requirements for the first year varied markedly between treatments, with implications for both emissions and patient experience. Omalizumab had the highest count, requiring two separate injections every two weeks to deliver a 225 mg dose. Reslizumab also had a high total count of 39 preparations and was the only regimen associated with significant API wastage. Each 210 mg dose of reslizumab is supplied using three 100 mg vials, leading to 90% of the third vial being discarded at every administration. In some countries, such as the UK, vial-based dosing of 200 mg is permitted.^36^ This reduces both the preparation count and emissions by a third compared with the 210 mg regimen. Aligning labelling and clinical practice to support standardised dosing, rather than weight-based dosing, offers an immediate opportunity to reduce waste and emissions.

Emerging long-acting biologics may also support alignment of reduced treatment frequency with lower environmental impact. Depemokimab, which was under regulatory review at the time of writing, is administered as a 100 mg dose twice yearly.^37^ At the non-fossil fuel electricity proportions applied in this analysis, the first-year emissions associated with depemokimab are estimated to be similar to benralizumab. While the per-patient carbon benefit is modest compared with existing low-emission options, the reduction in injections offers potential combined benefits for patient experience, healthcare resource use, and emissions.

Electricity source has been identified as a major contributor to the carbon footprint of biologic manufacture in several independent studies.^8,13,14,38^ In our analysis, differences in non-fossil fuel electricity (NFFE) proportions accounted for much of the variation between products, with manufacturer-reported values ranging from 22% to 91%. Modelling indicated that increasing NFFE to 100% could reduce first-year treatment emissions up to 90% for some products, and would reduce the difference between the therapies with the highest and lowest emissions by 91% to 17 kg CO2e per patient-year. Transitioning away from fossil fuel-derived electricity offers broad benefits for climate, health, and energy security, and is increasingly reflected in corporate and national decarbonisation commitments. For biologic manufacture, accelerating this shift represents a near-term opportunity to achieve major reductions in treatment emissions without changes to clinical practice.

Our study has several strengths. We applied a standardised approach that enabled direct comparison of the carbon footprint of different biologics. For emissions, we used MCF Classifier, which applies a consistent cradle-to-gate method across products. At the clinical level, we modelled the first year of treatment for a representative patient, ensuring that dose requirements and preparation counts were applied consistently across therapies. By integrating manufacturer-level electricity profiles, we were able to capture an important source of variation between products. To our knowledge, no other analysis has directly compared the carbon footprint of licensed biologics for severe asthma.

Our study also has important limitations. We used secondary data and a modelled approach rather than primary manufacturing data. Such data are not publicly available from manufacturers and, even if accessible, are not standardised in a way that would permit comparisons, which is central to our analysis. We applied manufacturer-level non-fossil fuel electricity (NFFE) proportions, as site- or product-specific values were not available. Given the influence of electricity sourcing on biologic emissions, this may have had a material impact on the results. Finally, we limited the scope of our analysis to cradle-to-gate boundaries rather than the full clinical pathway. Using a representative patient mitigates this for comparative purposes, but inclusion of downstream elements would provide a more complete picture of overall emissions.

Our findings show that biologic choice, dosing practices, and manufacturer electricity sourcing all have a material impact on the carbon footprint of severe asthma treatment. These factors present actionable opportunities for reducing emissions in the near term while maintaining quality of care. While our analysis focused on biologics for severe asthma, the same standardised approach can be applied to other therapies and therapeutic areas.

Embedding medicine-level carbon footprint data into prescribing, commissioning, and policy decisions offers a practical route to reducing the environmental impact of healthcare while maintaining high standards of patient care.

## Supporting information

Supplementary Material - Product Carbon Footprint Report (template)

## Contributors

HT and NR were jointly responsible for the concept, study design, method development, execution, analysis, writing, editing, and project administration. Both authors reviewed and approved the final manuscript. NR is the guarantor of the work.

## Funding

The development of MCF Classifier was funded by YewMaker. The application of the methodology to severe asthma biologics was supported by funding from ThermoFisher Scientific. ThermoFisher Scientific had no involvement in the study design, execution, analysis, interpretation of data, or in the writing or review of this manuscript.

## Competing Interests

HT and NR are employees of and own shares in YewMaker. NR is a Non-Executive Director of AstraZeneca.

## Patient and Public involvement

Patients and the public were not involved in the design, conduct, reporting, or dissemination plans of this research.

## Ethics approval

This study did not involve human participants or identifiable patient data and did not require ethical approval.

## Data availability statement

All data relevant to the study are included in the article or the supplementary material. Additional information is available from the corresponding author on reasonable request.

## Notes

### Competing Interest Statement

HT and NR are employees of and own shares in YewMaker. NR is a Non-Executive Director of AstraZeneca. YewMaker received funding from ThermoFisher Scientific for the application of the methodology to severe asthma. ThermoFisher Scientific had no role in study design, analysis, interpretation, or manuscript preparation.

### Funding Statement

The development of the MCF Classifier was funded by YewMaker. The application of methodology to severe asthma biologics was supported by funding from ThermoFisher Scientific. ThermoFisher Scientific had no involvement in the study design, execution, analysis, interpretation of data, or in the writing or review of this manuscript.

### Summary of Updates

Addition of ThermoFisher Scientific as a funder

