## Supplementary Material - Product Carbon Footprint Report (template) for "Comparative Analysis of the Carbon Footprint of Biologics for Severe Asthma"

### MCF Product Report

#### Template

(Input all items in red for the specific product)

---

#### Medicine: {Product} {API}
#### Dose: {Dose} mg
#### Form: {Volume} {Preparation format}
#### Manufacturer: {Manufacturer}

---

**Assessment Date:** {Month} {Year}
**Valid Until:** {Month} {Year}

*This study was conducted as an independent assessment compliant with GHG protocol, PAS 2050 and ISO 14067 product carbon footprint standards.*

### Table of Contents


**1. Acronyms and Abbreviations** **4**

**2. Goals of the Study** **5**

2.1. Statement of Compliance 5

2.2. Assurance of Independence 5

**3. Analysis Scope** **6**

3.1. Functional unit 6

3.2. Inventory type 6

3.3. Climate impacts 6

3.4. Time period 6

**4. System Boundaries** **7**

4.1. Process map 7

4.2. Attributable processes 7

4.2.1. API manufacture and formulation 7

4.2.2. Excipient manufacture 8

4.2.3. Packaging manufacture 8

4.3. Exclusions 8

4.3.1. Transport 8

4.3.2. Packaging 8

4.3.3. Greenhouse gas removals 8

**5. Life Cycle Inventory** **9**

5.1. Active pharmaceutical ingredients 9

5.2. Excipient ingredients 9

5.3. Packaging ingredients 9

**6. Methodology** **10**

6.1. API manufacture and formulation 10

6.2. Excipient manufacture 11

6.3. Packaging manufacture 12

6.4. MCF Ratings 12

6.5. MCF CarbonStamp™ 13

**7. Assessment of Data Quality** **15**

7.1. Technological representativeness 16

7.1.1. API manufacture and formulation 16

7.1.2. Excipient manufacture 16

7.1.3. Packaging manufacture 16
7.2. Geographical representativeness 16
7.3. Temporal representativeness 16
7.4. Completeness 16
7.5. Reliability 17
7.6. Uncertainty 17
**8. Results 18**
8.1. Carbon footprint per functional unit 18
8.2. Breakdown of carbon footprint by attributable process 18
8.3. Interpretation of results 18
8.3.1. API Manufacture and formulation 18
8.3.2. Excipient manufacture 18
8.3.3. Packaging Manufacture 18
8.4. MCF CarbonStamp™ 19
**9. Key References 20**

### 1. Acronyms and Abbreviations

**API**: Active Pharmaceutical Ingredient

**$CH_4$**: Methane

**$CO_2e$**: Carbon Dioxide Equivalent

**FDA**: Food and Drug Administration

**g**: grams

**GHG**: Greenhouse Gas

**GWP**: Global Warming Potential

**HFCs**: Hydrofluorocarbons

**INN**: International Nonproprietary Name

**IPCC**: Intergovernmental Panel on Climate Change

**ISO**: International Organization for Standardization

**kg**: kilograms

**mAb**: Monoclonal Antibody

**MCF**: Medicine Carbon Footprint

**$N_2O$**: Nitrous Oxide

**PAS**: Publicly Available Specification

**PFCs**: Perfluorocarbons

**$SF_6$**: Sulphur Hexafluoride

### 2. Goals of the Study

The goal of the study was to provide an independent cradle-to-gate life cycle assessment of the carbon footprint of **{Product} {Dose} mg in a {Volume} {Preparation format} manufactured by {Manufacturer}.**

This assessment was performed using Medicine Carbon Footprint (MCF) Classifier, a published, peer-reviewed, proprietary technology developed by YewMaker. MCF Classifier integrates modeled, empirical, and product-specific features. Product-specific features are taken from publicly available data sources to ensure method standardisation and independence.

The study was conducted as a third party independent assessment compliant with GHG protocol, PAS 2050, and ISO 14067 product carbon footprint standards.

#### 2.1. Statement of Compliance

*This study was conducted by YewMaker as an independent assessment, compliant with the GHG Protocol Product Life Cycle Accounting and Reporting Standard, ISO 14067:2018, and PAS 2050:2011.*

#### 2.2. Assurance of Independence

*This report was independently produced by YewMaker using its proprietary MCF Classifier methodology. Results are based on publicly available data and modelled estimates, with no input or influence from the manufacturer or funders.*

### 3. Analysis Scope

The analysis scope establishes the framework for the assessment, including the functional unit of measure, the inventory type, the climate impact categories, and the time period of the analysis.

#### 3.1. Functional unit

The functional unit of the analysis is a **{Volume} {Preparation format} containing {Dose} mg of {API} manufactured by {Manufacturer}**.

#### 3.2. Inventory type

The analysis is a cradle-to-gate inventory covering the life cycle of the product from raw material extraction until the final product leaves the manufacturer.

#### 3.3. Climate impacts

The analysis considers the carbon footprint of the product measured by Global Warming Potential (GWP) over a 100-year time horizon. The GWP is measured relative to carbon dioxide ($CO_2e$) as the standard unit. The results include emissions from all relevant greenhouse gases released during the product's life cycle, including but not limited to: carbon dioxide ($CO_2$), methane ($CH_4$), nitrous oxide ($N_2O$), sulphur hexafluoride ($SF_6$), perfluorocarbons (PFCs), and hydrofluorocarbons (HFCs).

#### 3.4. Time period

The analysis is based on the production of the functional unit in **{Year}** and was conducted in **{Month, Year}**. The results are valid for one year, until **{Month, Year}** reflecting the expected stability of the underlying data and assumptions.

### 4. System Boundaries

The system boundary defines the inclusions and exclusions for the cradle-to-gate life cycle assessment.

#### 4.1. Process map

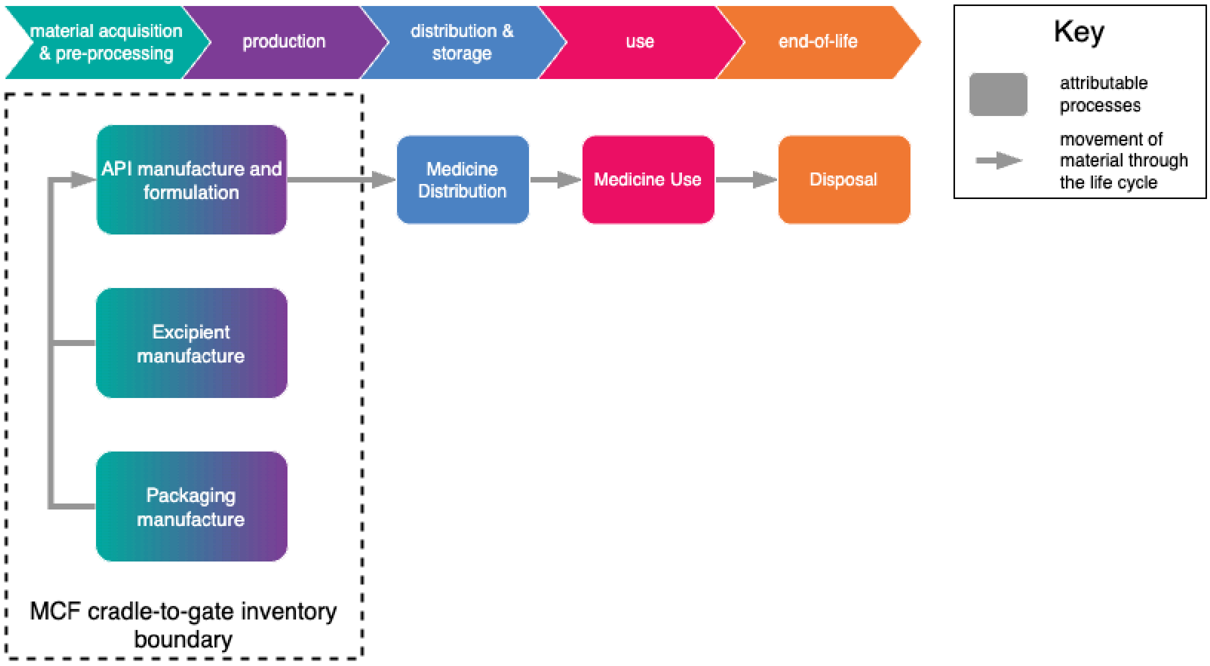


#### 4.2. Attributable processes

The carbon footprints of the following attributable processes were assessed:

1. Active pharmaceutical ingredient (API) manufacture and formulation
2. Excipient manufacture
3. Packaging manufacture

##### *4.2.1. API manufacture and formulation*

The included life cycle stages for API manufacture and formulation are:

1. Resource extraction
2. Raw materials received at the manufacturing site
3. Product formulation
4. Finished product leaving the manufacturer

###### *4.2.2. Excipient manufacture*

The included life cycle stages for excipient manufacture are:

1. Resource extraction
2. Raw materials received at the manufacturing site
3. Manufacture of excipients
4. Incorporation of excipients into the final product

###### *4.2.3. Packaging manufacture*

The included life cycle stages of packaging manufacture are:

1. Resource extraction
2. Raw materials received at the manufacturing site
3. Manufacture of primary packaging
4. Integration of primary packaging into the final product

##### 4.3. Exclusions

Certain processes were excluded from the analysis based on data availability and their minor contribution to overall emissions.

###### *4.3.1. Transport*

Transport processes were excluded due to the lack of representative secondary data and their minor contribution to the overall carbon footprint, as evidenced by previous life cycle assessments.

###### *4.3.2. Packaging*

The analysis includes primary packaging but excludes secondary and tertiary packaging.

###### *4.3.3. Greenhouse gas removals*

The analysis only includes emissions from the attributable processes. It does not account for greenhouse gas removals, such as those from offsetting or carbon capture.

### 5. Life Cycle Inventory

The life cycle inventory defines the ingredients assessed for each of the attributable processes.

#### 5.1. Active pharmaceutical ingredients

The active pharmaceutical ingredient is **{Dose} mg of {API}**.

#### 5.2. Excipient ingredients

The excipients are water and those listed below, which were collated from the product information sheet:

- Water
- {excipient}
- {excipient}
- etc

#### 5.3. Packaging ingredients

**{API} {Dose} mg** is supplied in a {**container type and volume**} with components and features as specified in the product information sheet.

#### 6. Methodology

The methodology used is MCF Classifier, a proprietary technology developed by YewMaker. MCF Classifier integrates modeled data, empirical values, and product-specific attributes to estimate the carbon footprint of medicines, leveraging data science, machine learning, literature review, and imputation to ensure a standardised and consistent assessment.

The methodology for each attributable process is detailed below.

##### 6.1. API manufacture and formulation

The carbon footprint attributable to API manufacture and formulation is calculated using a mass-based approach informed by scientific literature review, data science, and domain expertise. The method is based on the following key assumptions:

- Monoclonal antibodies (mAbs) are manufactured by cell culture fermentation of mammalian cells, a process that is highly energy-intensive.
- The carbon footprint is significantly influenced by the energy grid mix used during manufacturing, with high-carbon energy sources (fossil fuels) resulting in higher emissions compared to low-carbon energy sources (renewables and nuclear).
- The percentage of non-fossil fuel energy used globally by a manufacturer, as self-reported in publicly available information, is a reliable proxy for estimating the proportion of low-carbon energy used in the manufacturing process. This is referred to as the "global non-fossil energy penetration".
- A generalised method can be used for continuous and batch mAb production processes as global non-fossil energy penetration is consistent (for a given manufacturer) and differences in the processes are not known to significantly impact carbon emissions.

Using these assumptions the GWP (kg$CO_2$e per kg API) of mAbs manufacturing using different energy grid mixes ranging from 19-100% non-fossil fuel energy was estimated. A logarithmic curve was then applied to describe the relationship between GWP and non-fossil energy penetration, reflecting that increasing renewable or nuclear energy usage leads to a nonlinear reduction in GWP, as shown in the schematic below:

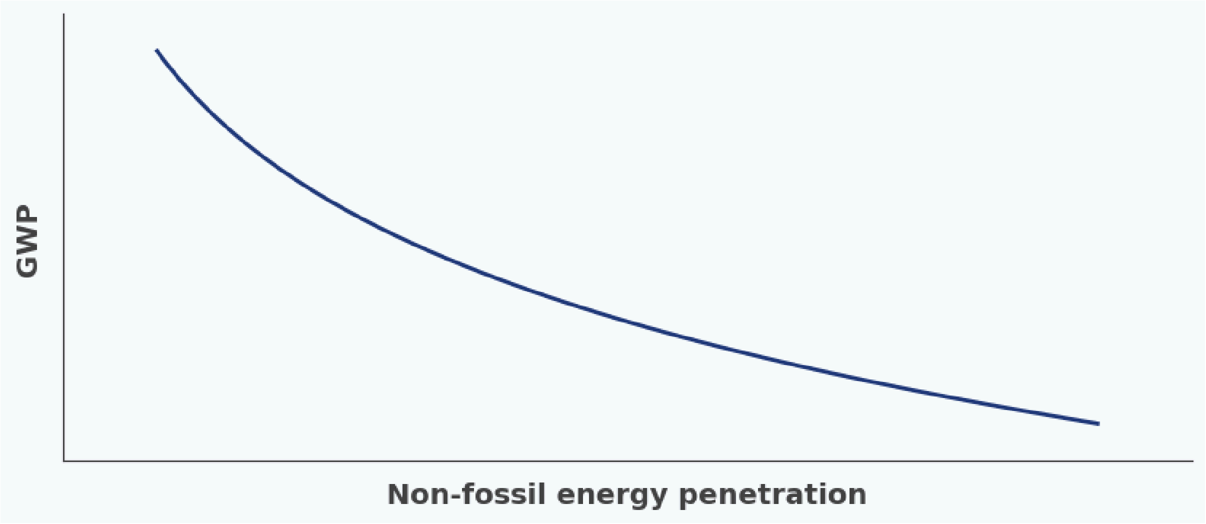


The calculated GWP value (kg$CO_2$e per kg API) is scaled to the functional unit by multiplying the GWP by the quantity of API in the functional unit.

#### 6.2. Excipient manufacture

The carbon footprint attributable to excipient manufacture is estimated using a mass-based approach informed by scientific literature review, data science, and domain expertise.

First, key excipients for the product class are identified based on their mass contribution, their GWP (kg$CO_2$e per kg produced), and their consistent presence across relevant products. Excipients that make a substantial contribution to the overall product mass and have a measurable GWP are prioritised in the analysis.

The GWP values for key excipients are obtained through a detailed literature search to identify relevant data and cradle-to-gate carbon emission values. Where specific data for an excipient is unavailable, the carbon footprint is imputed from similar excipients.

Next, product-specific excipients and their quantities (if available) are obtained from the Summary of Product Characteristics and/or the FDA Label. Adjustments to baseline excipient calculations for the product class are made based on this information, if required.

The final excipient carbon footprint is calculated as a weighted average of the individual footprints, considering their relative mass contributions and the typical (or known) total excipient mass in the product. Excipients that make a negligible (<1%) contribution to the total product carbon footprint are not considered.

These values are scaled to give a carbon footprint for the functional unit.

This methodology provides a practical and informed estimate of excipient carbon footprints.

#### 6.3. Packaging manufacture

The carbon footprint attributable to packaging manufacture is estimated using a mass-based approach informed by scientific literature review, data science. and domain expertise.

First, key packaging materials for the product class are identified based on their mass contribution and their GWP ($kgCO_2e$ per kg produced). Relevant data and cradle-to-gate carbon footprint values for these materials are obtained through a detailed literature review.

Next, product-specific packaging information is obtained from the Summary of Product Characteristics and/or the FDA Label. Adjustments to baseline calculations for the product class are made based on this information, if required.

**EITHER**:

This analysis considers the primary packaging in a **{Volume}** ml glass pre-filled syringe and is based on the following assumptions:

- All glass pre-filled syringes of the same size have a similar footprint, independent of manufacturer.
- The carbon footprint of packaging materials scales linearly with product volume at low volumes, such that a 2 ml pre-filled syringe is assumed to have double the carbon footprint of a 1 ml pre-filled syringe.
- The pre-filled syringe is designed for single-use only.

The final packaging carbon footprint is calculated based on the weighted contributions of the packaging materials, scaled to reflect the carbon footprint of a **{Volume}** ml glass pre-filled syringe.

**OR:**

This analysis considers the primary packaging in a **{Volume}** ml glass vial, and is based on the following assumptions:

- All glass vials of the same size have a similar footprint, independent of manufacturer.

- Vials of the same size have a similar footprint whether they are designed for multi-use or single-use.
- Any needles or syringes required to administer the product are part of the use phase and therefore not accounted for in the cradle-to-gate inventory.

The final packaging footprint for vials is calculated as the weighted contribution of the packaging materials, scaled to reflect the carbon footprint of a **{Volume}** ml glass vial.

This methodology provides a practical and informed estimate of packaging carbon footprints.

#### 6.4. MCF Ratings

To enhance usability, the carbon footprint ($gCO_2e$ per functional unit) is categorised using the MCF Ratings system. This traffic-light-style system groups carbon footprints into four categories based on a $\log_{10}$ scale as shown below:

| **MCF Rating** | **$gCO_2e$ per functional unit** |
|---|---|
| $CO_2$ Low | <10 |
| $CO_2$ Medium | ≥10-100 |
| $CO_2$ High | ≥100-1000 |
| $CO_2$ Very High | ≥1000 |

MCF Ratings simplify interpretation for stakeholders and facilitates comparisons across products and processes.

#### 6.5. MCF CarbonStamp™

The MCF CarbonStamp™ is a visual representation of the carbon footprint of the functional unit, designed to summarise the results in a concise and intuitive format. It provides both the precise carbon footprint value ($gCO_2e$) and the corresponding MCF Rating, allowing for quick interpretation and comparisons.

Key Features:

- Product identification: The assessed product, dose, and form that define the functional unit are displayed prominently. The brand name is used for an assessment of a specific product, otherwise the active pharmaceutical ingredient name is used.
- Carbon footprint value: The total $CO_2e$ emissions for the relevant functional unit are displayed centrally for clarity.
- Rating Ring scale: A color-coded ring surrounds the carbon footprint value to indicate the MCF Rating:
  - Low (Green)
  - Medium (Orange)
  - High (Red)
  - Very High (Purple)
- Interpretive visual: The Rating Ring is filled proportionally to the $CO_2e$ value, enabling a visual comparison across products at a glance.
- Validity Period: The time period for which the data is valid is indicated by a ‘valid until’ date and is based on the time period of validity specified in the corresponding Carbon Footprint Report.

The MCF CarbonStamp™ is included in the Results section of the Carbon Footprint Report as a standardised, standalone visual summary of the carbon footprint for the functional unit.

### 7. Assessment of Data Quality

Data quality is assessed using the qualitative framework set out in Chapter 8 of the GHG Product Life Cycle Accounting and Reporting Standard. The scoring criteria are based on Table 8.2 of the Standard as shown below and evaluate the following five indicators:

- Technological Representativeness
- Geographical Representiveness
- Temporal Representativeness
- Completeness
- Reliability

**Table [8.2] Sample scoring criteria for performing a qualitative data quality assessment**

| Score | Representativeness to the process in terms of: | | | | |
|---|---|---|---|---|---|
| | Technology | Time | Geography | Completeness | Reliability |
| **Very good** | Data generated using the same technology | Data with less than 3 years of difference | Data from the same area | Data from all relevant process sites over an adequate time period to even out normal fluctuations | Verified[4] data based on measurements[5] |
| **Good** | Data generated using a similar but different technology | Data with less than 6 years of difference | Data from a similar area | Data from more than 50 percent of sites for an adequate time period to even out normal fluctuations | Verified data partly based on assumptions or non-verified data based on measurements |
| **Fair** | Data generated using a different technology | Data with less than 10 years of difference | Data from a different area | Data from less than 50 percent of sites for an adequate time period to even out normal fluctuations or from more than 50 percent of sites but for shorter time period | Non-verified data partly based on assumptions or a qualified estimate (e.g., by sector expert) |
| **Poor** | Data where technology is unknown | Data with more than 10 years of difference or the age of the data are unknown | Data from an area that is unknown | Data from less than 50 percent of sites for shorter time period or representativeness is unknown | Non-qualified estimate |

Each indicator is scored qualitatively to evaluate the quality of data used in the carbon footprint assessment. The uncertainty of the data is also assessed.

When differences exist between attributable processes, individual assessments are presented. When data represent the entire life cycle, a single qualitative value is provided.

#### 7.1. Technological representativeness

##### *7.1.1. API manufacture and formulation*

The results for **{Product} {Dose} mg** production are calculated using a generalised method for monoclonal antibody production and the global energy grid mix of the manufacturer. This represents a "**Good**" qualitative GHG score.

##### *7.1.2. Excipient manufacture*

The results are calculated for a typical **{Preparation format}** incorporating data on the quantities and emissions of the specific excipients used in **{Product} {Dose} mg** formulation where they are available and different from a typical **{Preparation format}**. This represents a "**Good**" qualitative GHG score.

##### *7.1.3. Packaging manufacture*

The results are calculated for a typical **{Volume} {Preparation format}** incorporating data on specific materials used in **{Product} {Dose} mg** packaging where they are available and different from a typical **{Volume} {Preparation format}**. This represents a "**Good**" qualitative GHG score.

#### 7.2. Geographical representativeness

Geography is represented by including the global energy grid mix of **{Manufacturer}**. This represents a "**Good**" qualitative GHG score.

#### 7.3. Temporal representativeness

All calculations are based on data from 2021 onwards, reflecting recent manufacturing practices and energy grid data. This represents a "**Very Good**" qualitative GHG score.

#### 7.4. Completeness

The assessment encompasses the entire cradle-to-gate life cycle, including API manufacture and formulation, excipient manufacture and primary packaging. Minor exclusions, such as transport and secondary packaging are noted but do not significantly impact the results. This represents a "**Good**" qualitative GHG score.

#### 7.5. Reliability

The calculations are based on peer-reviewed, validated models and research data. While certain assumptions are necessary, these are supported by secondary data and domain expertise. This represents a “**Good**” qualitative GHG score.

#### 7.6. Uncertainty

GWP factors are considered with an uncertainty of ± 35 percent for the 90 percent confidence interval as indicated in the IPCC’s Fourth Assessment Report.

### 8. Results

#### 8.1. Carbon footprint per functional unit

- Functional unit: a **{Volume} {Preparation format}** containing **{Dose} mg** of **{API}** manufactured by **{Manufacturer}**
- Carbon Footprint: **{Carbon footprint}** $gCO_2e$
- MCF Rating: **{MCF Rating}**

#### 8.2. Breakdown of carbon footprint by attributable process

| Attributable process | Carbon Footprint ($gCO_2e$) | % Share |
|---|---|---|
| API manufacture and formulation | **{API emissions}** | **{API%}** |
| Excipient manufacture | ~0.00 | 0.0 |
| Packaging manufacture | **{Packaging emissions}** | **{Packaging%}** |

#### 8.3. Interpretation of results

##### *8.3.1. API Manufacture and formulation*

The majority of the carbon footprint (**{API%}**%) is attributed to API manufacture and formulation, driven by the energy-intensive processes involved in monoclonal antibody production.

##### *8.3.2. Excipient manufacture*

All excipients contribute negligibly (<1% of total product carbon footprint) and therefore the total is recorded as ~0.00 $gCO_2e$.

##### *8.3.3. Packaging Manufacture*

Packaging accounts for **{Packaging%}**% of the total carbon footprint, reflecting the emissions from the cradle-to-gate production of the **{Volume} {Preparation format}**.

#### 8.4. MCF CarbonStamp™

The MCF CarbonStamp™ provides a standalone visual summary of the results. It displays the following information:

- Product name, dose, and formulation: **{Product} {Dose}** mg **{Volume} {Preparation format}**.
- Carbon footprint value: **{Carbon footprint}** $gCO_2e$.
- MCF Rating: **{MCF Rating}** as represented by the {**Rating colour**} colour-coded Rating Ring.
- Visual Scale: The Rating Ring is filled proportionally to the $CO_2e$ value on a 100–1000 scale, allowing for quick visual interpretation of the product's environmental impact.
- Validity period: Valid until **{Validity period}**.

#### 9. Key References


1. Amasawa, E. et al. Correction to "cost–benefit analysis of monoclonal antibody cultivation scenarios in terms of life cycle environmental impact and operating cost" (2023). ACS Sustainable Chemistry & Engineering, 11(39), pp. 14645–14645. https://dx.doi.org/10.1021/acssuschemeng.3c05737
2. British Standards Institution PAS 2050:2011 - Specification for the assessment of the life cycle greenhouse gas emissions of goods and services. https://knowledge.bsigroup.com/products/specification-for-the-assessment-of-the-life-cycle-greenhouse-gas-emissions-of-goods-and-services?version=standard
3. Budzinski, K. et al. Streamlined life cycle assessment of single use technologies in biopharmaceutical manufacture (2022). New Biotechnology, 68, pp. 28–36. https://doi.org/10.1016/j.nbt.2022.01.002
4. Eckelman and Litan. Life Cycle Assessment of the Prefilled Apiject Injector (2024). https://apiject.com/wp-content/uploads/2024/10/ApiJect-Environmental-Study-Report-FINAL.pdf
5. European Medicines Agency. ANNEX I: Summary of Product Characteristics for **{Product}** INN **{API}** (**{Date}**). **{URL}**
6. FDA Label. **{Product}** Prescribing Information (**{Date}**). **{URL}**
7. Greenhouse Gas Protocol: Product Life Cycle Accounting and Reporting Standard. https://ghgprotocol.org/sites/default/files/standards/Product-Life-Cycle-Accounting-Reporting-Standard_041613.pdf
8. International Organization for Standardization (ISO) ISO 14067:2018 - Greenhouse gases — Carbon footprint of products — Requirements and guidelines for quantification. https://www.iso.org/standard/71206.html#:~:text=This%20document%20specifies%20principles%2C%20requirements,ISO%2014040%20and%20ISO%2014044
9. Intergovernmental Panel on Climate Change (IPCC) Fourth Assessment Report: https://archive.ipcc.ch/report/ar4/wg1/
10. IRENA. Renewable energy statistics 2024. International Renewable Energy Agency, Abu Dhabi.

https://www.irena.org/-/media/Files/IRENA/Agency/Publication/2024/Jul/IRENA_Renewable_Energy_Statistics_2024.pdf

11. Renteria Gamiz, A.G. et al. Environmental Sustainability Assessment of the manufacturing process of a biological active pharmaceutical ingredient (2019). Journal of Chemical Technology & Biotechnology, 94(6), pp. 1937–1944. https://doi.org/10.1002/jctb.5975
12. Taylor H, Mahamdallie S, Sawyer M, Rahman N. MCF classifier: Estimating, standardizing, and stratifying medicine carbon footprints, at scale (2024). Br J Clin Pharmacol. 2024; 90(11): 2713-2723. https://doi.org/10.1111/bcp.16229
